# Exploring the spread dynamics of COVID-19 in Morocco

**DOI:** 10.1101/2020.05.18.20106013

**Authors:** Mohamed Naji

**Affiliations:** University of Sidi Mohamed Ben Abdellah, Faculty of Sciences Dhar El-Mahraz, Laboratory of Applied Physics Informatics and Statistics (LPAIS), Fez, Morocco

## Abstract

Despite some similarities of the dynamic of COVID-19 spread in Morocco and other countries, the infection, recovery and death rates remain very variable. In this paper, we analyze the spread dynamics of COVID-19 in Morocco within a standard susceptible–exposed–infected–recovered–death (SEIRD) model. We have combined SEIRD model with a time–dependent infection rate function, to fit the real data of *i)* infection counts and *ii)* death rates due to COVID-19, for the period between March 2*^nd^* and Mai 15*^th^* 2020. By fitting the infection rate, SEIRD model placed the infection peak on 04/24/2020 and could reproduce it to a large extent on the expense of recovery and death rates. Fitting the SEIRD model to death rates gives rather satisfactory predictions with a maximum of infections on 04/06/2020. Regardless of the low peak position, the confirmed cases and transmission rate were well reproduced.

## 1. Introduction

The spread of an epidemic disease depends on the availability of susceptible individuals that can lead the virus to survive. New susceptible persons will keep transmission between individuals and therefore promote the virus to survive. As the severe acute respiratory disease COVID-19 presents a high transmission rate, it will very likely save it from been extinct, at least for several years. To slow-down the transmission of COVID-19 among the population and to avoid a high fatality rate, each country was forced to adopt drastic containment and preventive measures going from social distancing to a complete lock-down of the entire country.

Earlier in March 2020, Morocco found itself fighting against COVID-19, which seems to be one of the worst waves of transmissible diseases that hits the country in the past 200 years. Morocco took strict preventive measures to limit the spread of the virus, including the closure of borders, the nationwide lock-down, as well as the compulsory wearing of face-masks. Thanks to these measures, the COVID-19 spread seems to be controlled so far. However, a clear picture regarding the magnitude and the dynamics of the epidemic wave is still not yet drawn.

In this paper, we report the result of modeling the outbreak of COVID-19 in Morocco using SEIRD model, we discuss the validity and limits of the model and we give an estimate of the magnitude of the epidemic wave, *i.e*. number of infected individuals, transmission rate and an estimated end of the epidemic wave.

## 2. Methods and Theory

Although its novelty, the corona virus COVID-19 seems to follow the same spread dynamic like other pathogens. Since the outstanding work by W. O. Kermack and A. G. McKendrick [1] who laid the foundation of the SIR model (susceptible-infected-recovered), several mathematical models have been developed to describe the spread of several infectious diseases [2, 3, 4, 5, 6]. Nowadays, these models are not new and have many open-source Python and Matlab code implementations available online.

In this work, we have used a known mathematical model, which consists of five compartments *(S* − *E* − *I* −*R* −*D*): Susceptible (S), Exposed (E), Infected (I), Recovered (R) and Dead (D). Any individual in the fraction of the population that will get sick belongs to one of the aforementioned compartments. Since COVID-19 has an incubation time, the E compartment (for exposed) is allocated for individuals that have been infected but are not yet infectious themselves. The system of equations in the SEIRD model is given by:

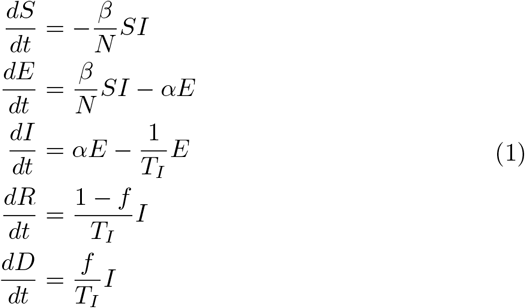

where *N* is the total number of individuals in the considered population. At each time *N* is equal to the sum of (Eq.1) *N* = *S* + *E* + *I* + *R* + *D*. *β* represents the infection rate, *i.e*., an expected amount of people an infected person infects per day, *α* is the incubation rate and *T_I_* is the average infectious period and *f* is the rate at which people die. The same model has been used elsewhere [7] to describe the widespread of COVID-19 in Italy. The total number of infected people in a population is determined by the *R*_0_ number. This can be expressed as the infection rate multiplied by the mean time of recovery or deaths. It describes the expected number of individuals an infected person infects in a susceptible population. Equivalently, *R*_0_ is computed as:

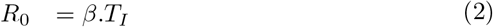

The system of Eq.(1) is solved with initial conditions [*S*(*t*_0_), *E*(*t*_0_), *I*(*t*_0_), *R*(*t*_0_), *D*(*t*_0_)] for some initial time *t*_0_. To test the model, we have used a set of parameters and initial conditions found in literature for the COVID-19 spread in China and Italy [*α* = 0.1, 1/*T_I_* = 2.73.10^−2^, *f* = 0.3 − 0.6], [*S*(*t*_0_) = *N* − 1, *E*(*t*_0_) = 1, 0, 0, 0] and let *β* float (data not shown). Although the model reproduces the gross features of the full time course of the data, it gives rather unrealistic values for the transmission rate *β*. The projection of COVID-19 dynamics in China and Italy to that of Morocco seems to be very rough, as the prevention measures taken by each country differ greatly. The latter should cause a decrease in the number of contacts between Infected and Susceptible population. Thus, *β* cannot remain constant or even abruptly jumps from one value to another. Therefore, we have adopted a time-dependent *β*(*t*) decreasing function of the form:

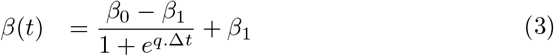

and reintegrated the system of Eq.(1) using the set of parameters shown in table 1.

**Table 1:** Table of fixed and best-fit parameters computed from the fitting routine as implemented in Python package.

| | $t_0$ | $\frac{1}{T_I}$ | $\alpha$ | $f$ | $q$ | $\beta_0$ | $\beta_1$ |
| --- | --- | --- | --- | --- | --- | --- | --- |
|  | days | (day<sup>-1</sup>) | (day<sup>-1</sup>) | (day<sup>-1</sup>) | (day<sup>-1</sup>) | (day<sup>-1</sup>) | (day<sup>-1</sup>) |
| **Fit to infection rate $I(t)$** | 7 | 0.25 | 0.25 | 0.03 | 0.11 | 0.51 | 0.19 |
| **Fit to death rate $D(t)$** | 7 | 0.25 | 0.25 | 0.03 | 4.99 | 0.62 | 0.15 |

## 3. Analysis and Discussion

### 3.1. Adjusting the Model to real data

#### 3.1.1. Daily infections I(t)

In order to come as close as possible to the real numbers and make informed predictions of the COVID-19 spread in Morocco, SEIRD model has been fitted to daily reported infection counts of COVID-19 data, for the period between March 2*^nd^* and Mai 15*^th^* 2020 (03/02/2020 – 05/15/2020), using the time–dependent *β*(*t*) function. To initialize the system of equations, the following initial parameters [*S*(*t*_0_) = *N* − 1, *E*(*t*_0_) = 1, 0, 0, 0] have been used, where *N*, represents the total number of population of Morocco and *t*_0_ corresponds to the first day of the first infected person. Morocco has detected the first infection on March 2*^nd^*. Thus, the infection could have in fact been days or weeks earlier. Consequently, we have tried several negative lag-time between 05 ≤ *t*_0_ ≤ 20.

The *t*_0_ value which seems to reproduce best the data was 7 days. Note that a lag-time of *t*_0_ = 30 days has been observed for Italy in order to reproduce the outset of the outbreak [10]. The best-fit parameters as well as those fixed are reported in table 1. Figure 1 (a) represents the best-fit to the daily counts of infections of COVID-19 in Morocco. It is noteworthy to visualize that the inflection point of *I*(*t*) was very well reproduced with our model, although it seems to underestimates the reported values around April 20*^th^* and Mai 8*^th^*. As the set of initial conditions were adequately adjusted, we believe that this mismatch is very likely due to the abrupt change in testing rate at this period related to the presence of several hotbeds of contagion throughout the country. One can also note that starting from March 8*^th^* a visible increase of the number of infections is also accompanied by an increasing number of the performed tests. Figure 2 (a) represents the predicted evolution of COVID-19 spread in Morocco, as calculated using the parameters set in table 1. It can be remarked from Figure 2 (a) that the global fit of the SEIRD model to the *I*(*t*) COVID-19 data in Morocco, while predicting the observed position of the epidemic peak, it does so at the price of a bad estimation of the growth of recovered and death population. Figure 3 (a) illustrates the evolution of the reproductive number *R*_0_(*t*) as function of time. The assessment of *R*_0_ = 2.05, looks reasonable at the early stage of the epidemic period, though it is much lower than that of COVID-19 worldwide. Such low predicted *R*_0_ would inevitably induce a massive decrease of the infected population. The total number of the cumulative confirmed cases (*C* = *R* + *D*) is also underestimated (see Figure 4), this suggests that the decay of the infection rate *I*(*t*) is higher than that recorded for the data. This discrepancy will very likely to get reduced when more data on the COVID-19 spread in Morocco will be available.

**Figure 1:**
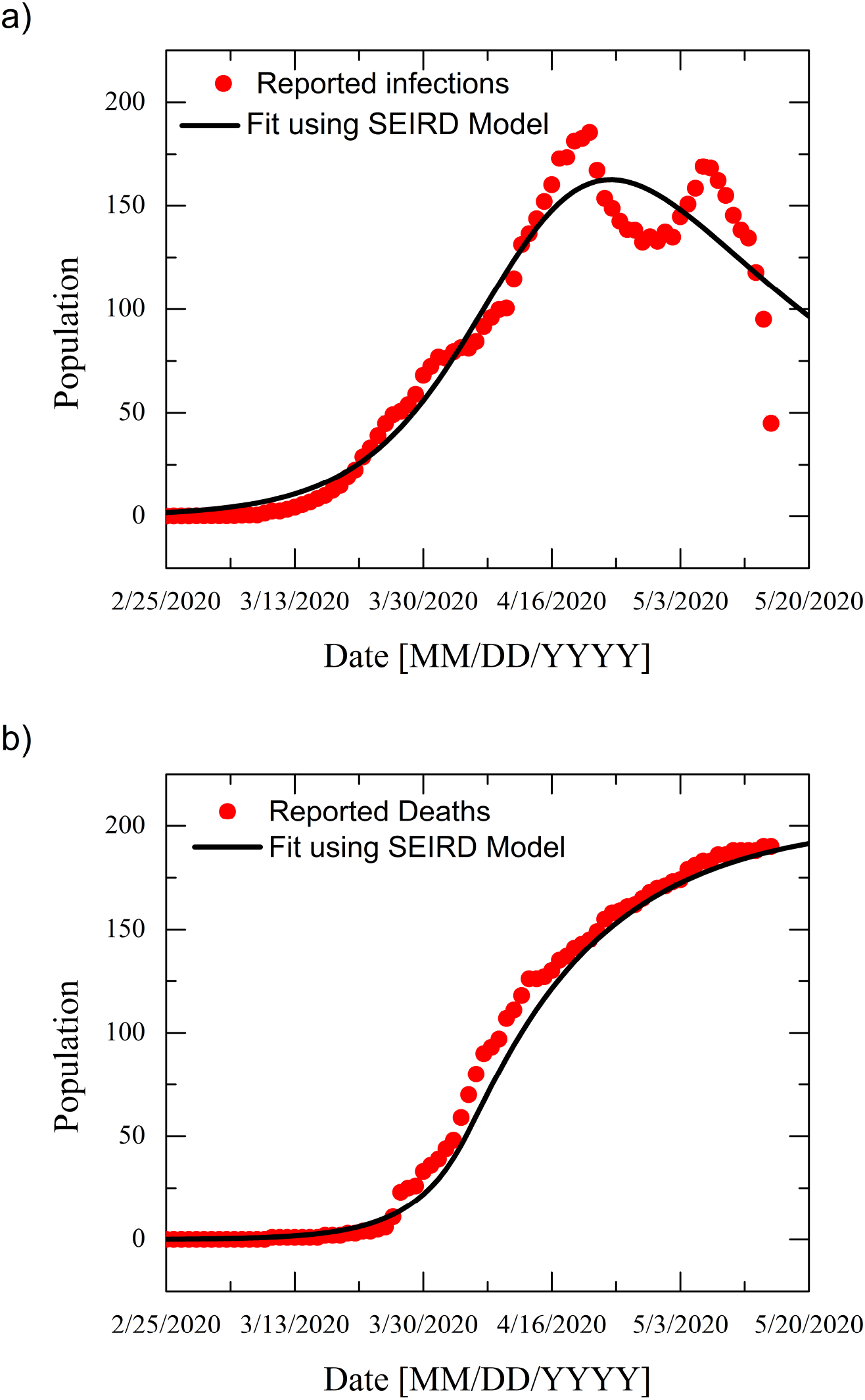
Evolution of the COVID-19 epidemic spread in Morocco. (a) Fit of COVID-19 daily infection counts in Morocco. (b) Fit of COVID-19 death cases in Morocco. Symbols represent the data obtained from the repository of the Center for Systems Science and Engineering (CSSE) at Johns Hopkins University (JHU) [8, 9], solid lines correspond to best-fit of the SEIRD model.

**Figure 2:**
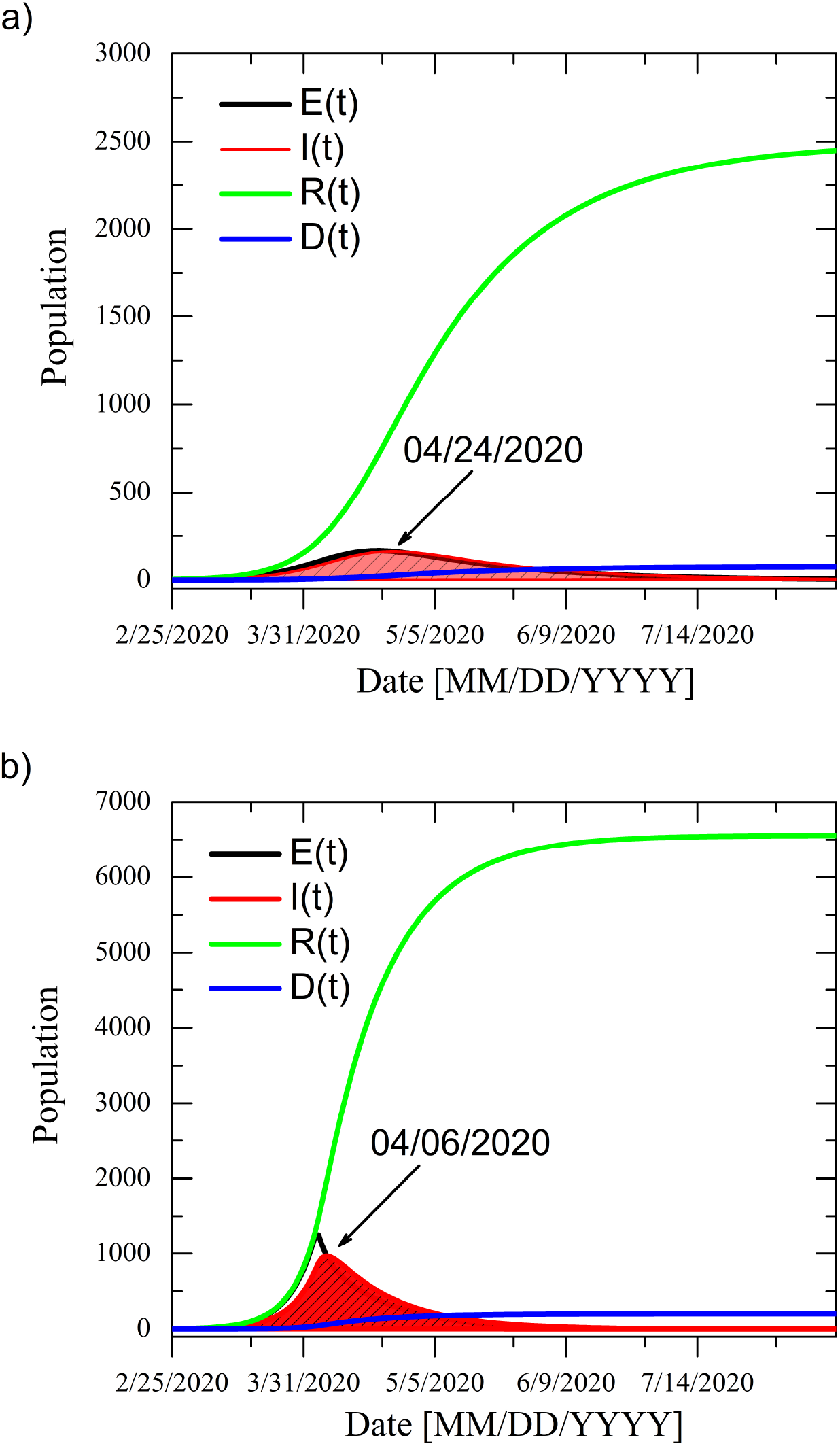
Prediction of the spread dynamics of COVID-19 in Morocco as calculated through SEIRD model. (a) Prediction based on fitting the infection counts. (b) Prediction based on fitting the death cases in Morocco. Arrows indicate the peaks of infection.

**Figure 3:**
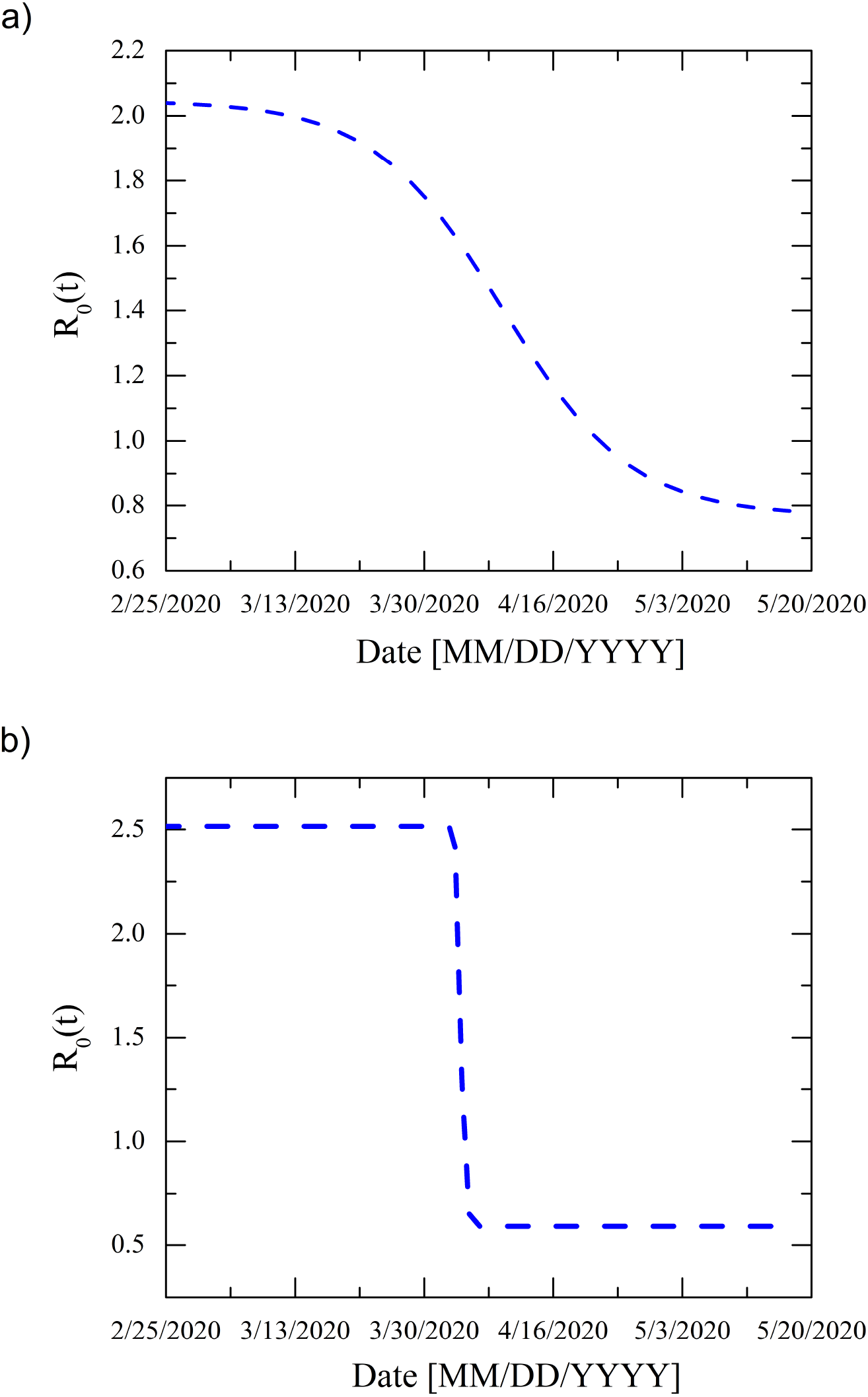
Prediction of *R*_0_ number in Morocco. (a) Prediction based on fitting the infection counts. (b) Prediction based on fitting the death cases in Morocco.

**Figure 4:**
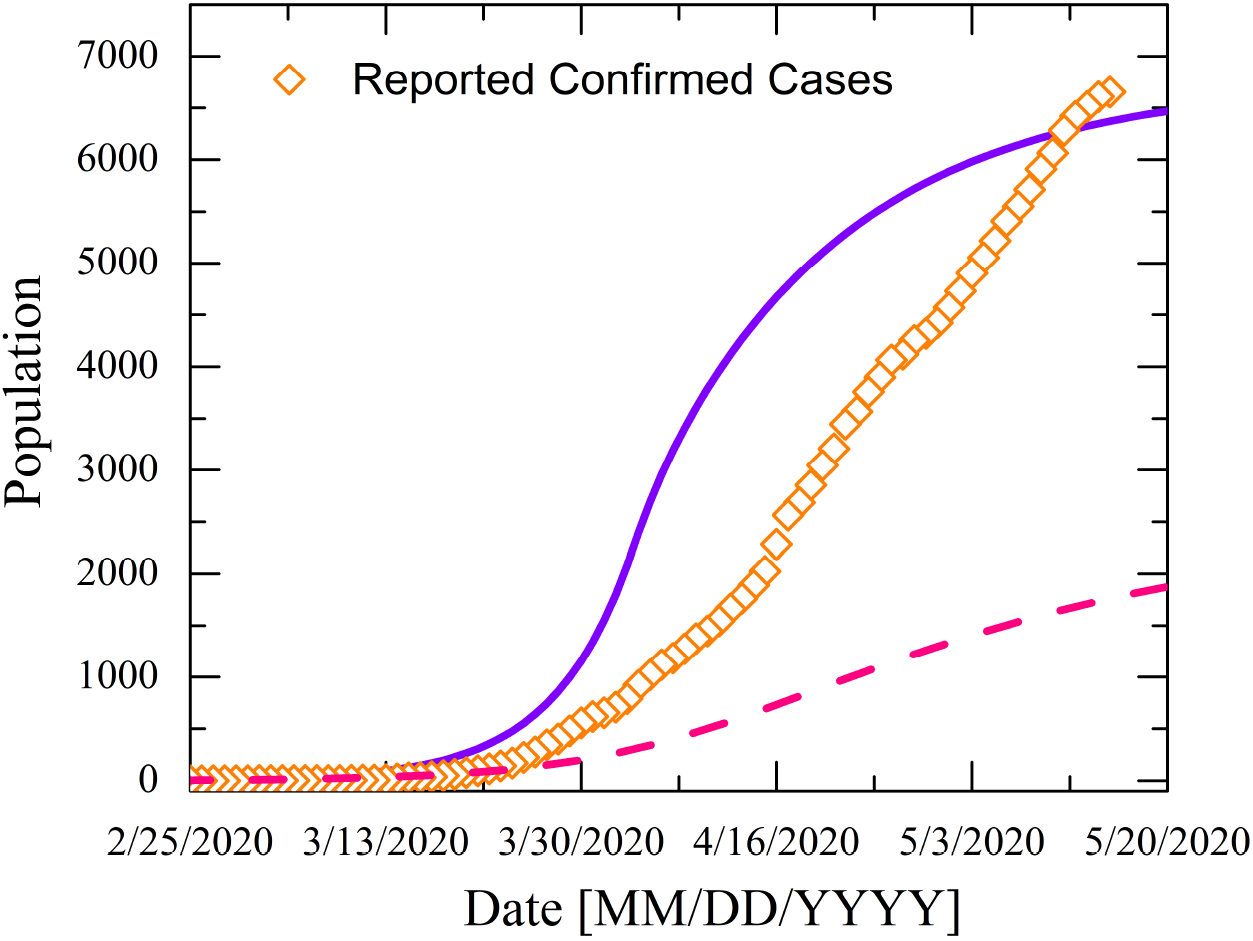
Evolution of the COVID-19 Confirmed cases in Morocco. *(Dashed-line)* prediction of confirmed cases based on fitting the infection counts. *(Diamond symbols)* prediction of confirmed cases based on fitting the death rate. *(Solid-line)* represent the data obtained from CSSE at Johns Hopkins University (JHU) [8, 9].

Despite, its reproducibility of the infection dynamics, the analysis of the COVID-19 outbreak in Morocco, using the fit of SEIRD model seems to be very crude. First, the data contain several spikes and looks noisy (though it has been averaged), and second the number of tests that have been performed, is very low, as compared to those performed in Italy, Germany or China. Note that during the early stage of the epidemic period, Morocco made tests to only those who presented COVID-19 symptoms, so obviously, infected individuals who did not present symptoms, have escaped to the tests. This explains the low estimation of confirmed cases.

#### 3.1.2. Death rate D(t)

In order to avoid external factors and get more accurate analysis of the COVID-19 spread in Morocco, SEIRD model has been fitted death cases due to COVID-19 in Morocco, for the period between 03/02/2020 – 05/15/2020. As Morocco, did not have an intensive wave of death cases due to COVID-19, as the one observed in Italy or China for example, the reported COVID-19 death cases should be reliable as it is hard to miss any case.

To initialize the system of equations, we have used the same initial parameters [*S*(*t*_0_) = *N* − 1, *E*(*t*_0_) = 1, 0, 0, 0] with *t*_0_ = 7. The best-fit parameters as well as those fixed are reported in table 1. Figure 1 (b) represents the best-fit to the reported death cases due to COVID-19 in Morocco. The fit gives rather satisfactory reproduction of the expected “S” shape of the death data. Further, it reaches a plateau of about 220 death cases, which looks quite reasonable, if considering the low number of individuals in intensive care and the low fatality rate ≃ 0.03 in Morocco. This suggests also that our fixed initial conditions were very well estimated. Figure 2 (b) represents the predicted evolution of COVID-19 outbreak in Morocco, as calculated using the parameters set in table 1. As it is seen from Figure 2 (b), the position of the epidemic peak, occurs 18 days earlier than the one reported in real data and also the number of the infected individuals is ten times higher than that in the real case. Analysis of *R*_0_ = 2.5 at the early stage of the epidemic period seems be rather satisfactory as it agrees very well with that computed from the real reported infection data (Figure 3). This also explains the low infection cases and low fatality rate seen in Morocco. It is important to note that starting from 03/30/2020, *R*_0_(*t*) decreased considerably as a result of preventive measures together with complete nation lock-down, adopted by the Moroccan Government on 03/20/2020 (just 10 days later).

Indeed, the flaw of the model in predicting the infection rate will undoubtedly get reduced when more data on the COVID-19 outbreak in Morocco will be available but one can already make some remarks. The down-shift of the epidemic peak as well as the predicted large number of infected cases can be understood if we take into account the initial low testing performed by Morocco, especially at the early stage of the epidemic period. Theoretically, by increasing the testing rate, we can expect a down-shift of the inflection point and an increase of infected cases, as this population would be detected earlier. One more important evidence about the robustness of this model is the total number of confirmed cases *C*(*t*) which saturates at ≃ 7000. Figure 4 shows the evolution of the predicted confirmed cases *C*(*t*) together with real COVID-19 data. Even if the predicted curve is down shifted (as consequence of the inflection point in the predicted infection curve), both curves seems to follow the same trend, with a constant difference (due always to the same reasons discussed above). The predicted saturation value saturates at the end of the epidemic wave (≃ 7000), seems to be realistic with the trend of COVID-19 dynamics in Morocco, unless other external factors enter into account such as hotbeds of contagion.

## 4. Conclusion

In this paper, we have analyzed the data of COVID-19 epidemic wave that hits the Moroccan territory starting from March 2^nd^. Thanks to the aggressiveness of containment measures, the country has recorded one of the low death rates across the Globe. The analysis of COVID-19 data using a standard model composed of five compartments: susceptible, exposed, infected, recovered and dead (SEIRD) during the period 03/02/2020 – 05/15/2020 suggest that the country has already gone through the peak of infections with a minimum of infection is expected around 06/14/2020. The fit of SEIRD model to the COVID-19 infection counts in Morocco, placed the infection peak on 04/24/2020 and could reproduce to large extent the daily number of infections. However, it underestimates the recovery and death rates, leading thus to a low prediction of the total number of the confirmed cases. The *R*_0_ = 2.05 at the beginning of the epidemic period is lower than that expected for COVID-19 worldwide. Even if the model seems to be reliable in predicting the infection rates, it does so at the expense of other compartment of the population. Accordingly, the interpretation of such predictions should be take with care. On the other hand, fitting the SEIRD model to death rates due to COVID-19, looks more reliable especially as the data did not depend on external factors. Indeed, the death rate depends on several factors, like the health system, the average age of the elderly people, their health conditions, availability of intensive care units, etc. But in the absence of statistics regarding the factors influencing death rates, it is still premature to draw conclusions. Besides that, the fit gives rather satisfactory reproduction of the death data. Further, it reaches a plateau of about 220 death cases, which seems reasonable, with the data reported so far. The drawback of such approach is the down-shift of the position of the epidemic peak and the large number of infections reported on that day. Although, this is rather harsh but it still not very surprising if taking into account the low testing rate. Moreover, the predicted *R*_0_ and the total number of infections *C*(*t*) further support the robustness of this approach when fitting the model to data.

## Data Availability

All data used were obtained form Covid-19 data repository at github.
URL https://github.com/CSSEGISandData/COVID-19

